# Optimal Control Strategy Applied to Dynamic Model of Drug Abuse Incident for Reducing Its Adverse Effects

**DOI:** 10.1101/2020.05.02.20088468

**Authors:** Md. Azmir Ibne Islam, Md. Haider Ali Biswas

**Author notes:** Mathematics Subject Classification (MSC 2010): 34A12, 34A34, 34H05, 35B35, 49K15, 93A30, 92D30.

## Abstract

The present world is facing a devastating reality as drug abuse prevails in every corner of a society. The progress of a country is obstructed due to the excessive practice of taking drugs by the young generation. Like other countries, Bangladesh is also facing this dreadful situation. The multiple use of drug substances leads an individual to a sorrowful destination and for this reason, the natural behavior of human mind is disrupted. An addicted individual may regain his normal life by proper monitoring and treatment. The objective of this study is to analyze a mathematical model on the dynamics of drug abuse in the perspective of Bangladesh and reduce the harmful consequences with effective control policies using the idea of optimal control theory. The model has been solved analytically introducing a specific optimal goal. Numerical simulations have also been performed to review the behaviors of analytical findings. The analytical results have been verified with the numerical simulations. The analysis of this paper shows that it is possible to control drug addiction if there is less interaction among general people with the addicted individuals. Family based care, proper medical treatment, awareness and educational programs can be the most effective ways to reduce the adverse effects of drug addiction in a shortest possible time.

## 1. Introduction

Drug addiction has become a major problem nowadays. The present world is experiencing the devastating effects of drug abuse day by day. It works as a hindrance to present civilization. It can also restrain us from accomplishing our goals or dreams of life. Generally, the use of substances that are injurious to health is known as drug abuse. It is believed that substance abuse induces sleep or produces excitement or insensibility which has a negative effect on the mental and physical health of an individual. Addiction is not only confined with one single cause but also a number of factors (genetic, brain chemistry, environmental issues and psychological behaviors) are working together behind the practice of consuming drugs. As addiction can trap anyone, people are greatly influenced by the social factors around themselves. The developments in human and national economic growth are about to decline as consuming of illicit drug abuse is increasing rapidly. Drugs have very harmful consequences both for human body and mind. Drug addiction is detrimental to health, even sometimes leads consumers to death. Also the structure and function of brain are greatly hampered due to seeking drugs randomly. As drug addiction can decrease self-control and human ability to make sound decisions, the world is now undergoing a big challenge to meet this crisis. But it can be managed effectively like other chronic and infectious diseases. Fortunately, there are treatments that can help people to counteract with the disruptive effects of addiction and regain control of lives. Research shows that treatment in rehabilitation centers ensures a better life for the patients who are habitual drug addicts [30]. An addict can achieve a sustained recovery in his or her future life without drugs after getting treatment from rehabilitation centers. Drug addiction is significantly gaining attention in whole world and increasing different kinds of social and public health problems nowadays. Drug addiction can be controlled by raising public awareness among the addicts, introducing voluntary activities and enforcing laws against the dealers.

Bangladesh has achieved a significant success in socioeconomic development in recent years. Diverse challenges have been taken to increase the life expectancy of people, reduce population growth, increase educational facilities and improve the health of maternal and child. Though the country is advancing with its valuable efforts, another problem has recently arisen which is working as an impediment towards those developments. Such specific problem that becomes the major threat for our country is known as ‘drug addiction’. This human devastation agent has spread its steps to every nook and corner across the country. Bangladesh, though not a drug producing country, is vulnerable for drug abuse because of its geographical location [1].

The tendency of drug addiction can mostly be seen among the young generations who are the leaders of future world. As drug addiction is increasing rapidly throughout the country, the youngsters are getting closure to addiction more and more, ruining their valuable lives and tending the country to a blind future. Many factors such as family problem, social factors, financial and legal issues, school or college environments, physical inability, failure of romance, job stress, poor self-esteem, high level sports competition, loneliness, low cost and easy access to drugs and local trafficking zone etc. are acting behind this worst situation.

Mathematical modeling has provided a quantitative insight into diverse fields. Several models have been published in recent years considering various problems regarding practical life. The literature and development of mathematical epidemiology are well documented and can be found in [3, 5, 6, 14, 28, 42, 44, 51]. Mathematical modeling has also launched its steps to explain different social problems and find out the alternative ways for reducing those problems. On the other hand, optimal control theory is another useful part of mathematics that describes the control policies [4, 7, 8, 12, 27, 40, 48, 52, 53]. Mathematical Model and Optimal Control are closely related to each other [40]. Optimal Control deals with the problem of finding a control law for a system so that a certain optimality criterion is achieved by maximizing or minimizing a particular cost function. The formulation of an optimal control problem requires a mathematical model to be controlled. In short, a mathematical model works as a framework for an optimal control problem [40].

Drug addiction has become an important issue in recent years as it ruins the future of young generation. A model on heroin epidemics describes the global behaviour with distributed time delays. Heroin is fairly responsible for the transmission of human immunodeficiency virus (HIV) and hepatitis C virus in China [6, 41]. The impact and adverse effects of an illicit drug use have been analyzed and further extended with two optimal strategies. The objective of the first optimal approach is to reduce the social influence between the susceptible and drug users, while the second aim is to increase the treatment facilities in rehabilitation [48]. A perspective, based on the principles of mathematical epidemiology, shows the consequences of opiate addiction in Europe specifically in Ireland. Individuals, who have a habitual tendency to use opiate drug, are responsible for the destruction of their own physical fitness and mental abilities and also put their families into a great trouble. Later it has been realized that prevention is indeed better than cure [67]. Computational models on the basis of neurobiology of drug abuse have been formulated with the help of mathematical modeling in order to illustrate the potential contribution due to addiction [2]. A model correlated with drug abuse has recently showed that the repeated use of drug substance is merely responsible for the increase of infectious diseases around the world [58] while on the other hand, a tobacco model has been established to obtain the periodic motion from tobacco addiction with complete recovery, relapse rate and addiction stages [3].

In this paper, we have developed a model on the consequences of drug abuse. The desire of this paper is to discover the fact that how the tendency of drug seeking mentality is prevailing among the susceptibles and continuing its chain to covert an individual into a light and heavy addict. We have also established the optimal control policy for minimizing drug addiction so that the young generation may become conscious about the adverse consequences of this deadly drug substance. The valuable efforts of rehabilitation centers can play a vital role in controlling yaba addiction by giving proper treatment to the addicted fellows. The main aim of our present paper is to limit the use of drugs, reduce and minimize the harmful effects of drug abuse as well as take an effective effort to maintain an addiction free environment.

The specific objectives of this study are to propose a mathematical model on the dynamics of drug abuse and formulate an optimal control strategy. As drug addiction is increasing rapidly throughout the country, some effective plans are needed for reducing the harmful consequences. In this stage, our model will a play a significant role and can also motivate the general people. Our main purpose is to increase consciousness among young generation about the adverse effects of drug addiction, raise public awareness and minimize the harmful consequences so that this unfavorable situation can be controlled.

## 2. Model Formulation

A population size *N*(*t*) is subdivided into five compartments: susceptible compartment *S*(*t*) (individuals who are not drug users, but at a high risk of taking drugs), light drug users *L*(*t*), heavy drug users *H*(*t*), drug users under treatment in rehabilitation R(t) and quitter population Q(t) (individuals who are fully determined not to take drug substances anymore). The schematic diagram of the model is shown in Figure 2.1.

**Figure 2.1.**
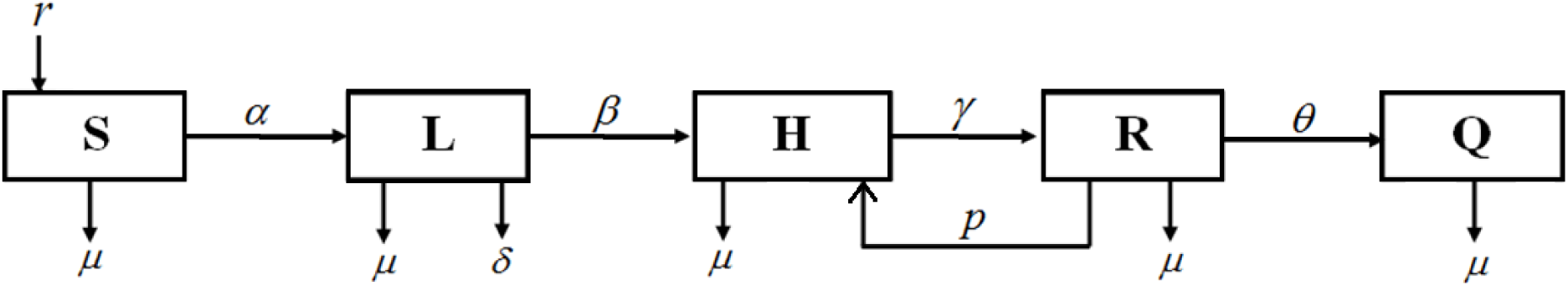
Compartmental representation of drug addiction model

Taking the Figure 2.1 into consideration, the model can be represented by the following system of non-linear ordinary differential equations:

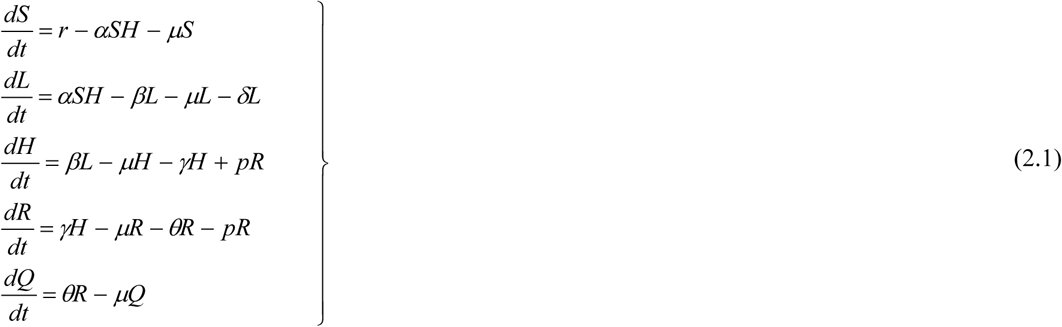

where, *S*(0) > 0,L(0) ≥ 0,*H*(0) ≥ 0,R(0) ≥ 0,Q(0) ≥ 0 and *N = S + L + H + R + Q*.

In the above model, the parameter *r* represents the recruitment rate of population, *µ* is the natural mortality rate, *α* is the interaction rate among the susceptibles and light drug users whereas *β* is the effective rate at which light users convert into heavy drug users. The constant *δ* represents the removal rate from addiction without treatment and *γ* is the rate at which heavy addicts are being sent to rehabilitation for treatment. In real life, despite getting treatment from rehabilitation centres, some addicts may again become a heavy drug user. Therefore, it is assumed that *p* is the relapsing rate for which a drug addict under treatment in rehabilitation converts into a heavy user again. Finally, *θ* is the fully recovered rate from addiction.

## 3. Formulation of Optimal Control Model

In this section, the drug abuse model has been analyzed newly by introducing an objective function using the well-known optimal control theory. The model is extended to observe the behaviors in the presence of control measures. We have used three control policies in order to minimize the number of drug addicted population. Then the model has been analyzed with several optimality conditions and numerical simulations have also been performed which show an effective outcome that we have expected as our assumption. The schematic diagram of model (2.1) can be reformulated as optimal control form as shown in Figure 3.1.

**Figure 3.1.**
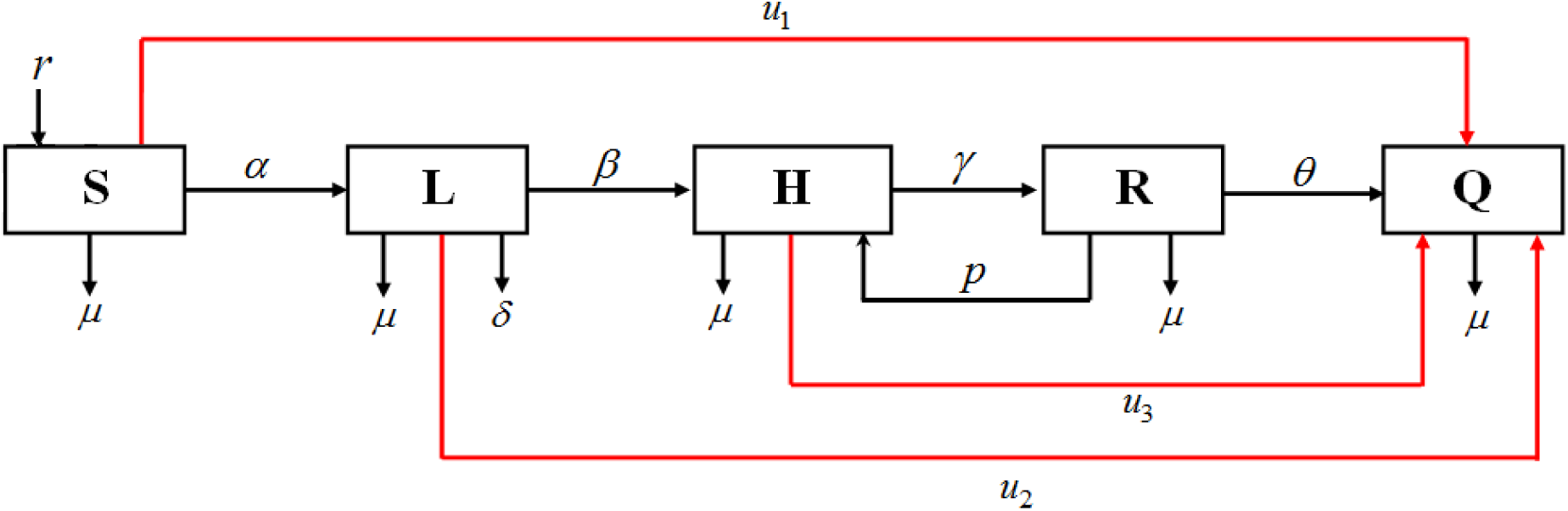
Compartmental representation of drug addiction model with control

Three control variables *u*_1_, *u*_2_, *u*_3_ have been introduced where *u*_1_ is the awareness and educational programs, *u*_2_ is the family based care and *u*_3_ represents the effectiveness of rehabilitation centers. It is to be mentioned that when the effectiveness of rehabilitation centers increases, the relapse case decreases. After considering these three control policies, the model (2.1) can be written in optimal control problem as

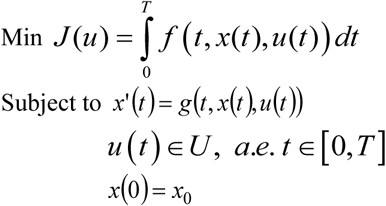

Here the objective functional is

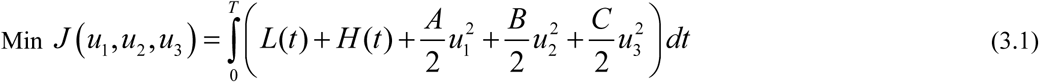

where, *A*, *B* and *C* are the weight parameters balancing the costs for reducing the addicted individuals. The dynamic constraints can be reformulated as

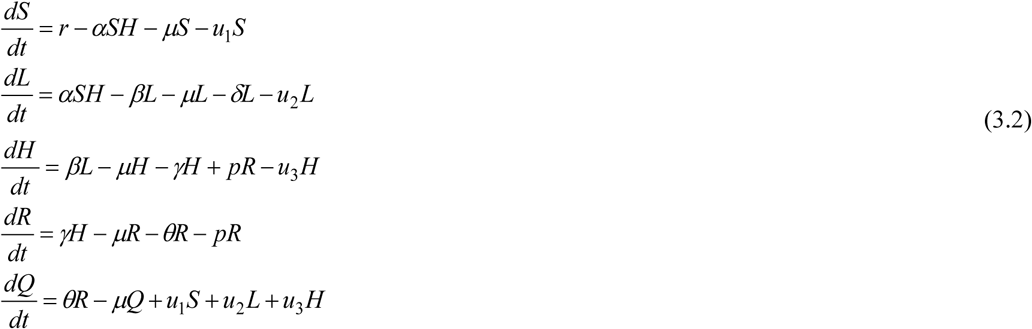

with the initial conditions, *S*(0) = *S*_0_, *L*(0) = *L*_0_, *H*(0) = *H*_0_, *R*(0) = *R*_0_, *Q*(0) = *Q*_0_

Equation (3.1) is the Lagrange type objective functional, where 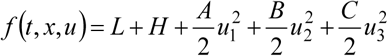 and *g*(*t*, *x*, *u*) is the right side of the system of differential equations (2.1) updated with control variables i.e. *x*(*t*) = (*S*(*t*), *L*(*t*), *H* (*t*), *R*(*t*), *Q*(*t*)), *u* (*t*) = (*u*_1_(t), *u*_2_(t), *u*_3_(t)) and *g* (*t*, *x*, *u*) = (S′, *L*′, *H*′, *R*′, *Q*′) In other words, the system of differential equations (3.2) along with the objective functional (3.1) describe the controlled model.

Now, we have to find a set for optimal control strategies 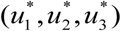 such that

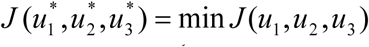

where, the control set 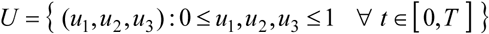

## 4. The necessary and sufficient conditions of optimal control

In this section, we are going to study the sufficient condition for the existence of an optimal control of the system of ordinary differential equations (3.1) [23]. Later on, Pontryagin’s Maximum Principle is used to characterize the optimal control functions and derive the necessary conditions for our control problem [40, 55].

### 4.1. The existence of an optimal control

In this subsection, the existence of an optimal control for the model (3.1) will be examined. In order to establish the existence of an optimal control and the uniqueness of the optimality system, the boundedness of solutions to the system (2.1) for a finite time interval is needed. We begin by examining the priori boundedness of the state solutions of model (2.1). For the model system (2.1) to be epidemiologically meaningful, all the state variables should be nonnegative for all time. We need to show that the total population is bounded for all *t* ≥ 0.

From model (2.1), the rate of change of total population is

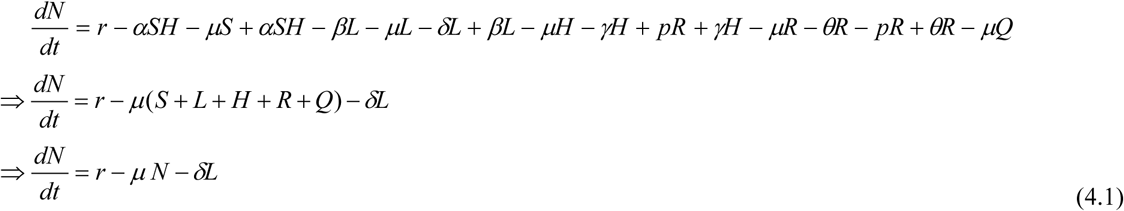

In the absence of drug addiction, we obtain

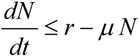

Now solving this, we have

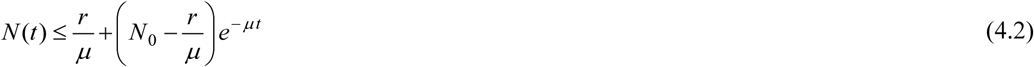

From (4.2), it is clear that the total population *N*(*t*) will approach the threshold 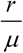 as *t* → *∞*. This implies that if the initial total population *N*_0_ is less than 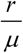 i.e. if 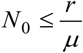 then 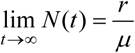. So, definitely 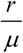 is the upper bound of N.

On the other hand, if 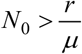, then *N*(*t*) will decrease to 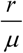 as *t* → *∞*. This means that if 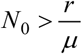 then the solution (*S*(*t*), *L*(*t*), *H*(*t*), *R*(*t*), *Q*(*t*)) enters the region Ω or approaches it asymptotically.

So it can be concluded that the region Ω is positively invariant under the flow induced by the model (2.1). Therefore, the model is both mathematically and epidemiologically well-posed in the region Ω. It is therefore sufficient to study the dynamics of the model (2.1) in Ω.

#### Theorem 4.1.

There exists an optimal control 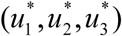 to problem (3.1).

##### Proof.

To prove this theorem, the concept described in [23] has been followed.

Considering 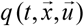 as the right-hand side of constraint, we would like to satisfy the following conditions:

1. *q* is of class C^1^ and there exists a constant *c* such that

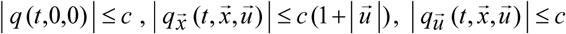
2. The admissible set Q of all solutions to system (3.2) with corresponding control in *U* is nonempty.
3. 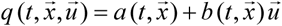
4. The control set *U =* [0,1] × [0,1] × [0,1] is closed, convex and compact.
5. The integrand of the objective functional is convex in U.

To verify the above conditions, we have

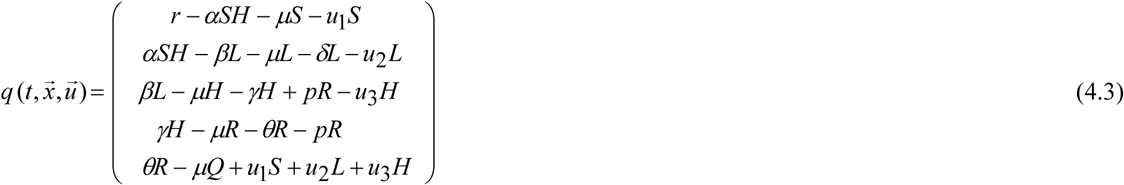

Therefore 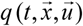 is of class C^1^ and 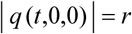.

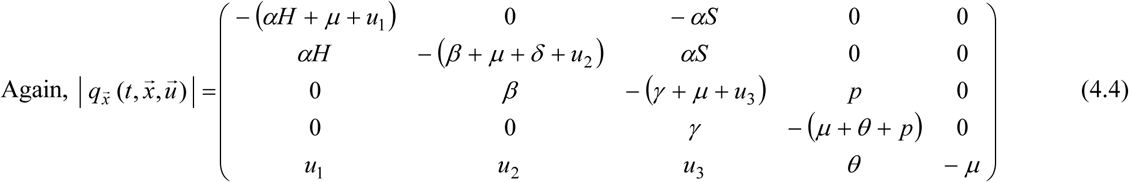

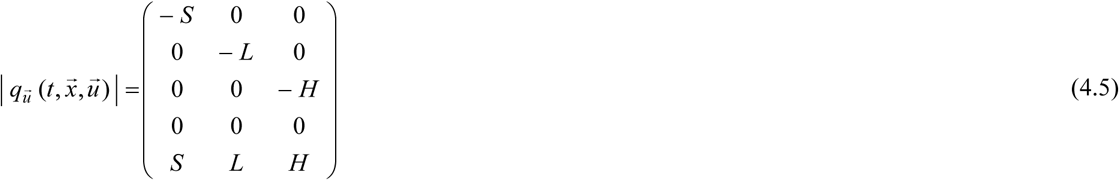

Since *S*(*t*), *L*(*t*), *H*(*t*), *R*(*t*), *Q*(*t*) are bounded, there exists a constant *c* such that

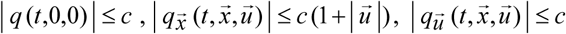

So, condition (1) is satisfied.

From condition (1), it is easily visible that there exists a unique solution to the system (3.2) for a constant control, which further implies that condition (2) holds.

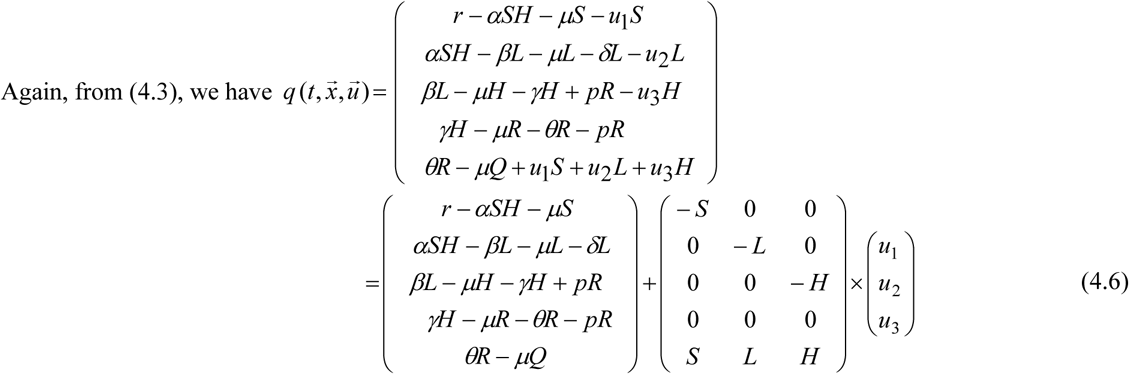

therefore, condition (3) also holds.

Condition (4) is obvious from the definition and the proof of this theorem can be completed by verifying condition (5). In order to show the convexity of the integrand in the objective functional 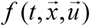, we have to prove that

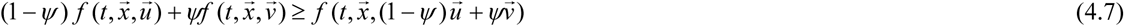

where, 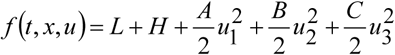 and 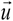 and 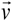 are two control vectors with *ψ* ϵ [0,1]. It follows that

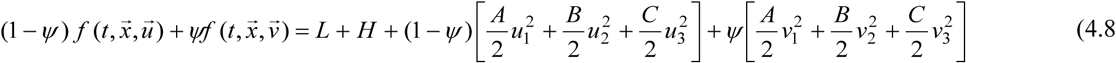

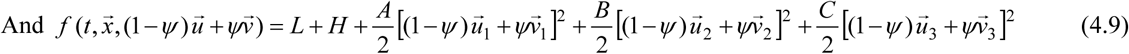

Furthermore, we have 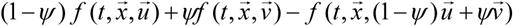

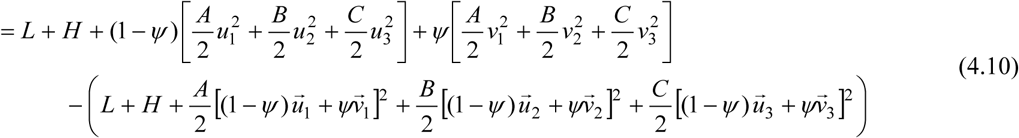

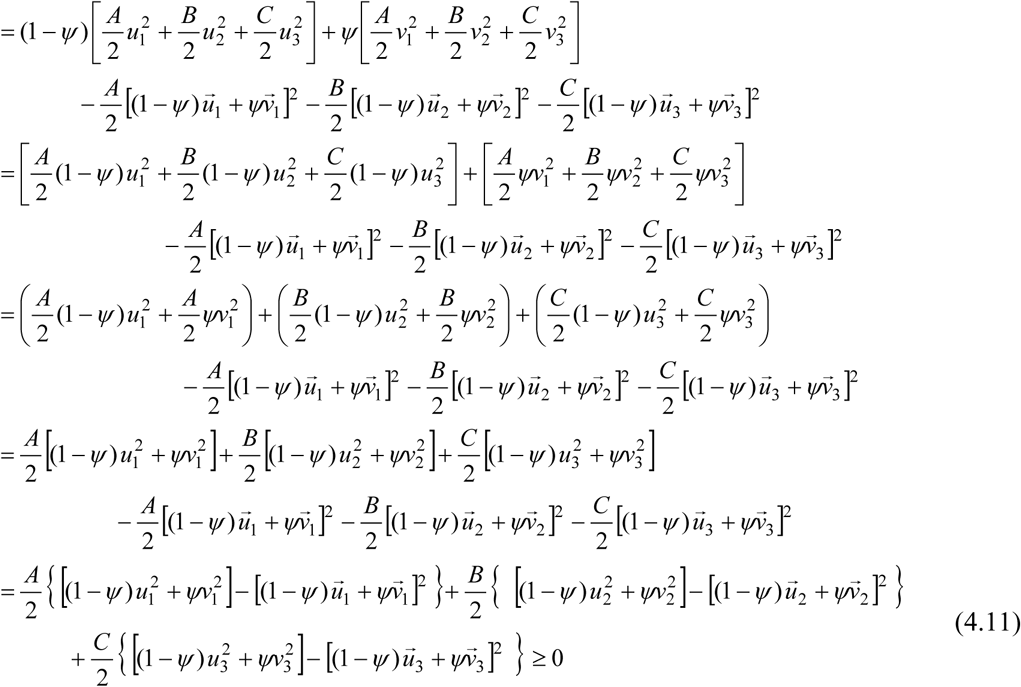

Finally, this is the complete proof.

### 4.2. Characterization of an optimal control

Since there is an optimal control for the objective function (3.1) subject to the model system (3.2), our target is to derive the necessary conditions for this optimal control [40, 55].

Using Pontryagin’s Maximum Principle, the Hamiltonian function *H_m_* w.r.t (*u*_1_,*u*_2_,*u*_3_) is

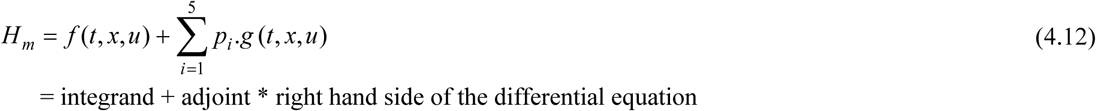

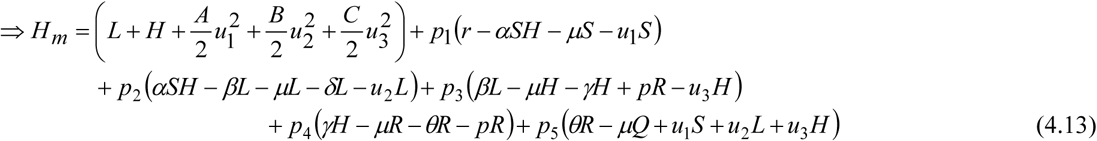

where, the variables *p_i_*, *i =* 1, 2, 3, 4 and 5 are adjoint or co-state variables.

The optimality system of equations can be obtained by considering the appropriate partial derivatives of Hamiltonian (4.13) with respect to the associated state variables. The following theorem is a consequence of the maximum principle.

#### Theorem 4.2.

For an optimal control pair 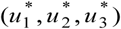 and corresponding solutions to the state system *S*^*^*, L*^*^*, H*^*^*, R*^*^*, Q*^*^ that minimize the objective functional (3.1),
there exists adjoint variables *p_i_*, *i =* 1, 2, 3, 4 and 5, satisfying the following adjoint equations:

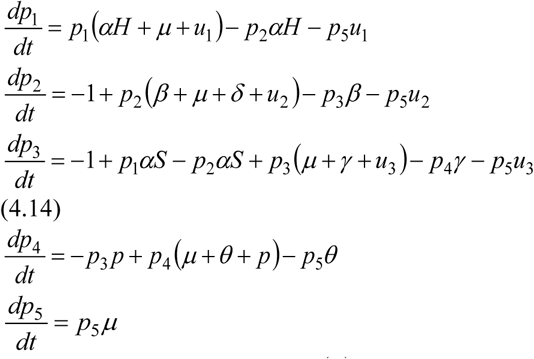

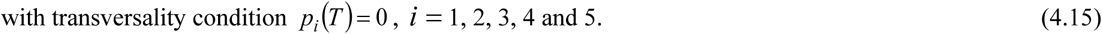

Again, the optimal pair may be characterized by the piecewise continuous functions

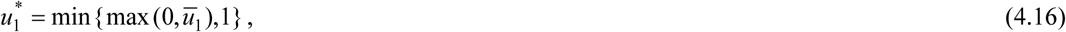

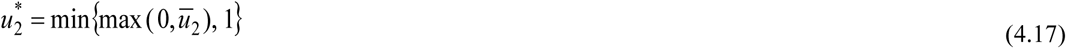

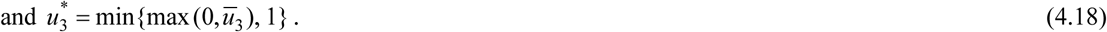

where,

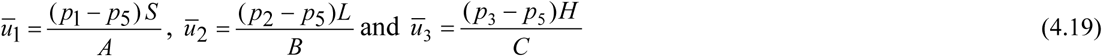

and *p_i_*, *i =* 1, 2, 3, 4 and 5 are the solutions of the adjoint system (4.14).

##### Proof.

Considering 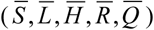 be the optimum values (*S*(*t*)*,L*(*t*)*,H*(*t*)*,R*(*t*)*,Q*(*t*)), the adjoint system (4.14) is obtained directly from the application of Pontryagin’s Maximum principle [40, 55]. The differential equations governing the adjoint variables are obtained by differentiating the Hamiltonian function. 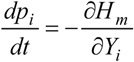
where, *Y_i_* =[*S*, *L*, *H*, *R*, *Q*]and *i=*1, 2, 3, 4 and 5.
Or,

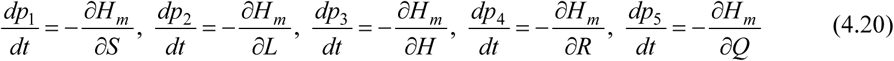

and *p_i_*(*T*) = 0, *i =* 1, 2, 3, 4 and 5.

We simply differentiate the Hamiltonian *H_m_* with respect to the controls (*u*_1_,*u*_2_,*u*_3_) at the optimal control functions and then equate the resulting expressions to zero,

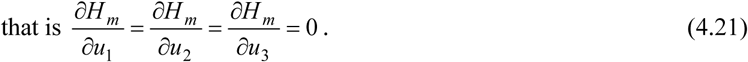

Solving for 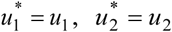 and 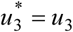, we derive the optimal controls 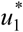, 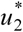 and 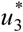 as given by the Theorem 4.1.

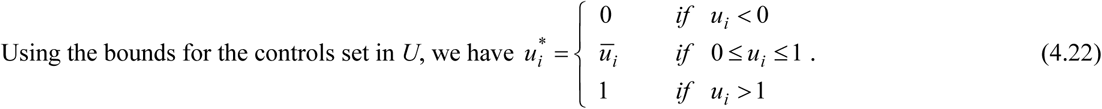

Finally, the optimality system consists of the state system (3.2) with the initial conditions, adjoint system (4.14) with transversality conditions (4.15) and optimality condition (4.16) - (4.18).

Thus, we have the following optimality system:

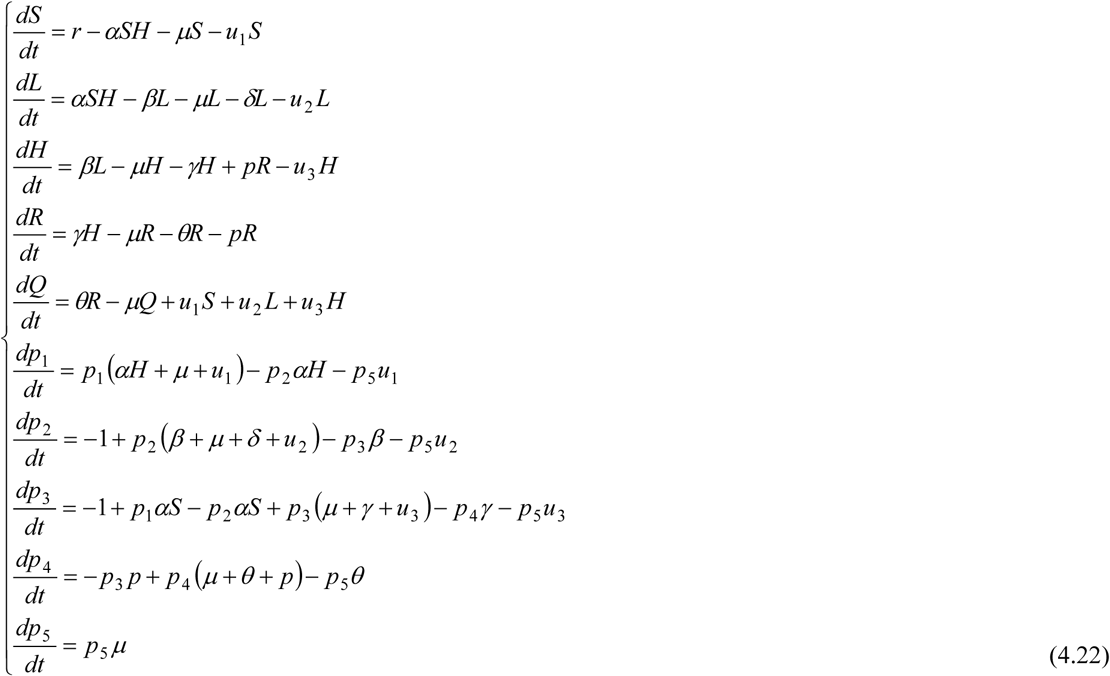

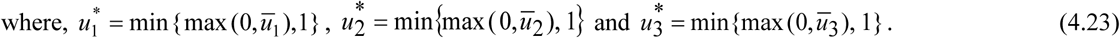

### 4.3. Uniqueness of the optimality system

Due to the boundedness for the state system, the adjoint system has bounded coefficients which is linear in each of the adjoint variables. Therefore, the adjoint system has finite upper bounds. The following proposition is needed for the proof of the uniqueness of the optimality system for the small time window.

#### Proposition 4.1.

The function *u*^*^(m) = min{ max{*m*, *a*},*b* }which is Lipschitz continuous in *m*, with *a < b* for some fixed nonnegative constants [34].

Now, we are about to prove the uniqueness of the optimality system in the same way as described in [25, 34, 71].

#### Theorem 4.3.

The solution to the optimality system (4.22) is unique for sufficiently small *T*.

##### Proof.

The proof can be done by contradiction.

That means, the boundedness for the state system and the adjoint system, the Lipschitz continuity of optimal control functions and some elementary inequalities play key roles to reach the contradiction.

Considering (*S*, *L*, *H*, *R*, *Q*, *p*_1_, *p*_2_, *p*_3_,*p*_4_,*p*_5_)and 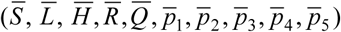 are the two non-identical solutions to the optimal system (4.22).

In order to show that the two solutions are equivalent, it is convenient to make a change of variables.

Let,

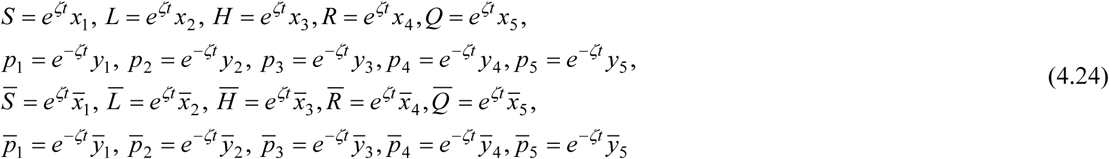

where, *ζ* is a positive constant to be chosen later.

After introducing the new variables, the optimality conditions can be converted to

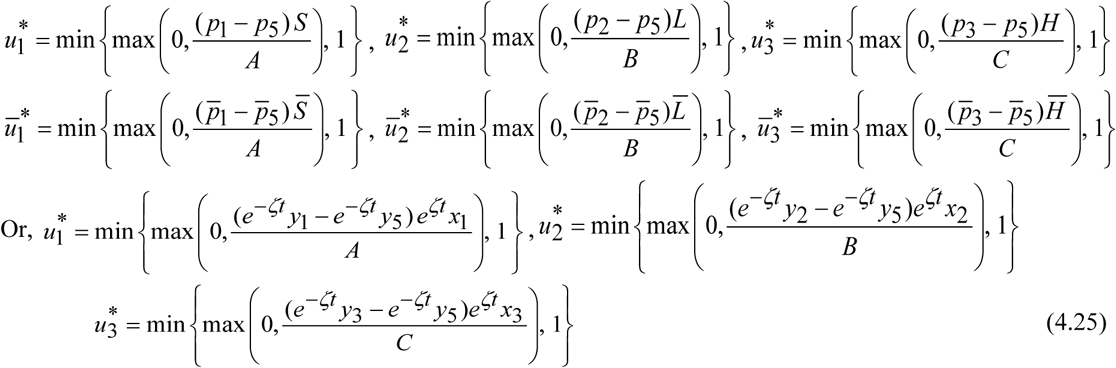

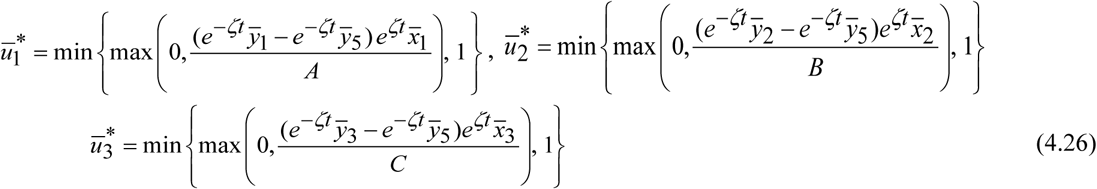

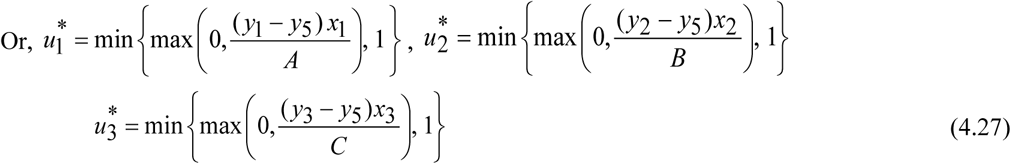

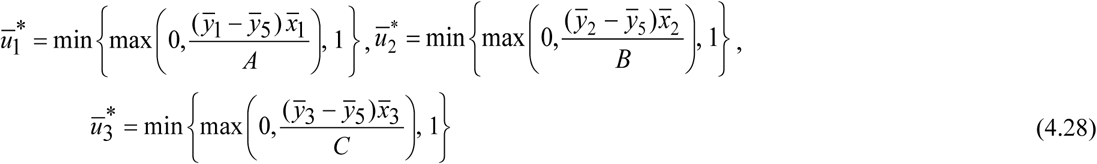

For simplicity of notation, we generally ignore the dependence on time in the following except in the case that a specific time is intended. Now, the difference of the resulting equations for *x_i_*, 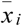, *y_i_* and 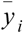 has been considered.

From the first equation of (4.22), we get

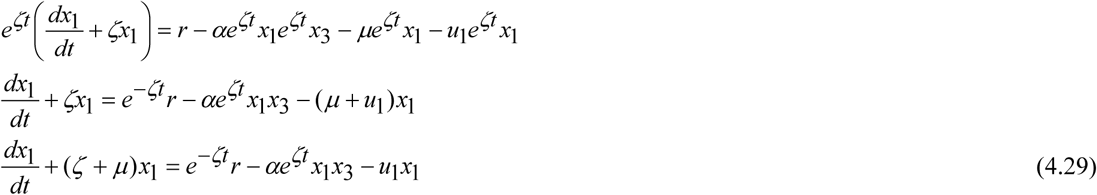

Similarly,

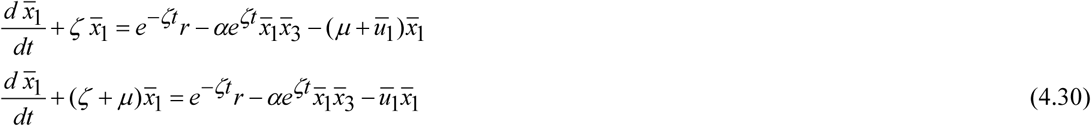

It follows from the above two equations that

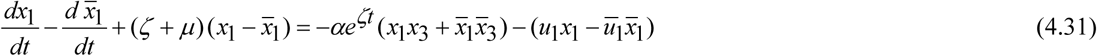

Multiplying the equation (4.31) by 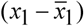 and then integrating both sides from 0 to *T*, we obtain

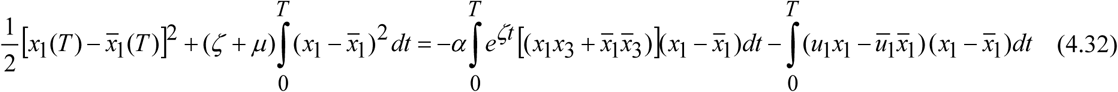

In order to simplify the right-hand expressions of (4.32), we need some elementary inequalities.

By the elementary inequality (*a* + *b*)^2^ ≤ 2(*a*^2^ + *b*^2^), we have

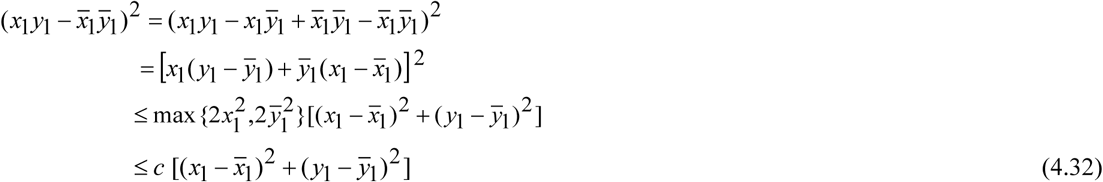

where, *c* depends on bounds for *x*_1_, 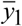.

Another inequality is also needed to obtain the specific estimation: If 0 ≤ *b* ≤ *a*, then 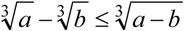.

Using **Proposition 4.1** and the above inequalities, we have the following estimation of 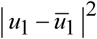. We are dividing our estimation in five cases as follows.

Considering the cases:

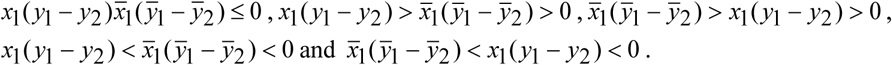

We finally can conclude

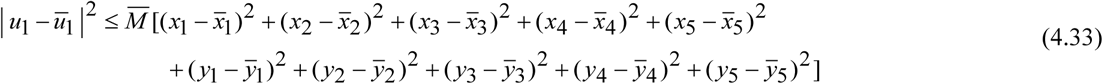

where, 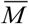 depends on bounds for 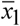, *y*_1_, *y*_2_.

Based on the above arguments, we find

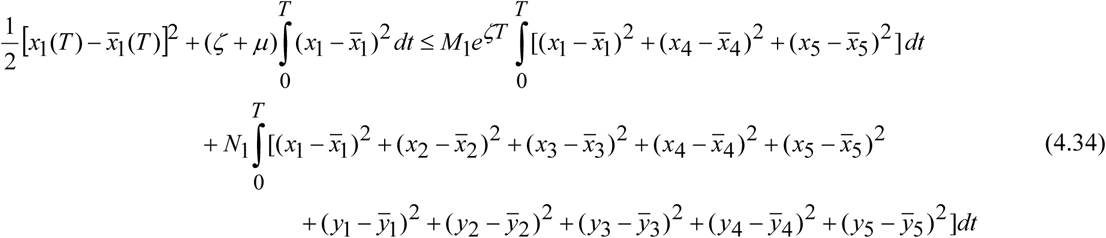

where, *M*_1_, *N*_1_ are the appropriate upper bounds. Similarly, we can obtain the following inequalities for 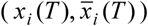 and 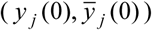 with *i* = 2,3,4,5 and *j* =1,2,3,4,5:

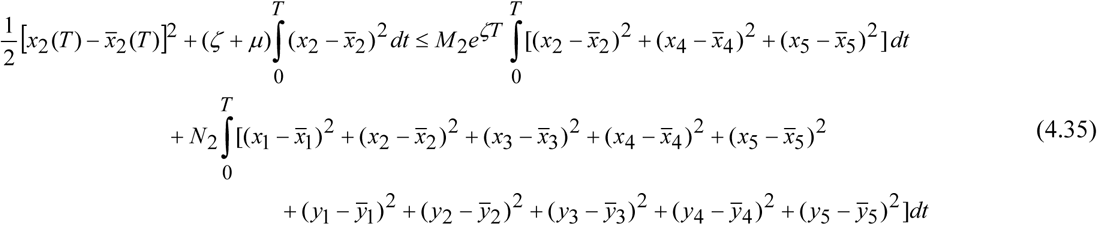

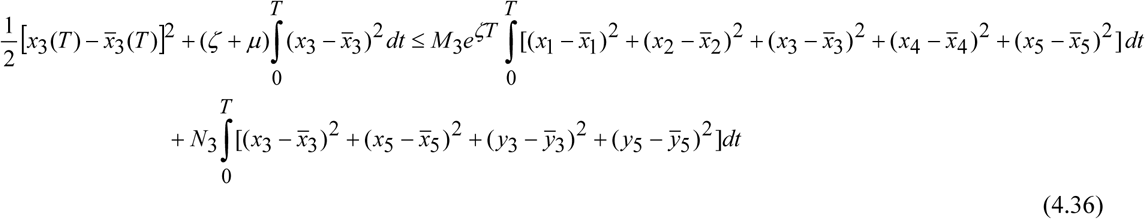

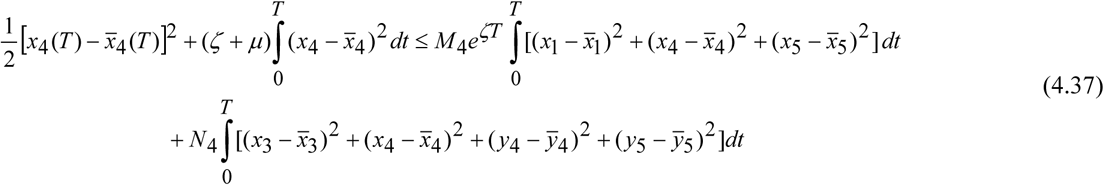

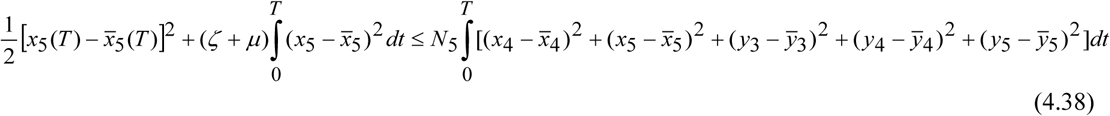

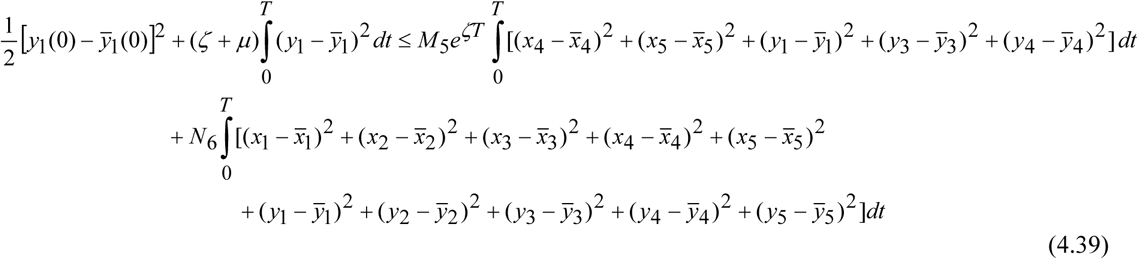

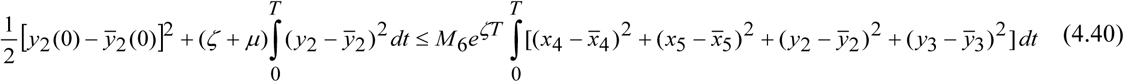

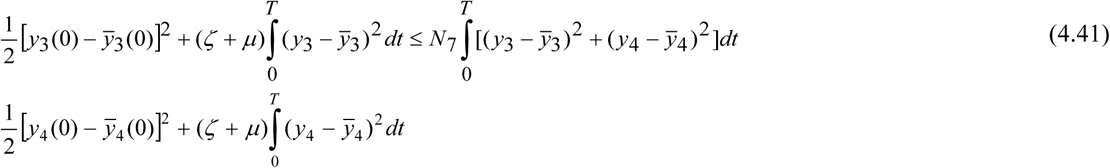

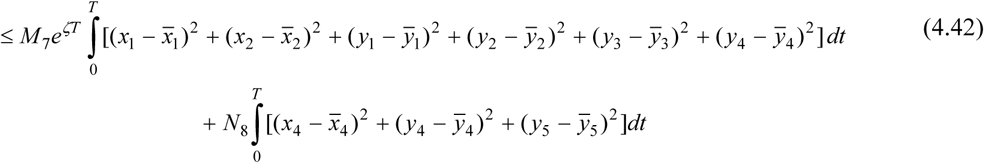

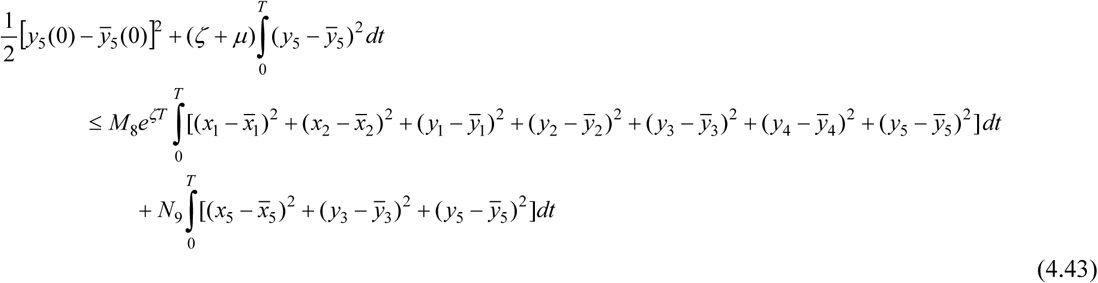

where, *M_i_* (*i =* 1,2,…,8)and *N_j_* (*j =* 1,2,…,9) depend upon the coefficients and the bounds of the state variables and co-state variables.

It can easily be seen that the coefficients of each integral term is nonnegative for sufficiently small *T*. This implies that

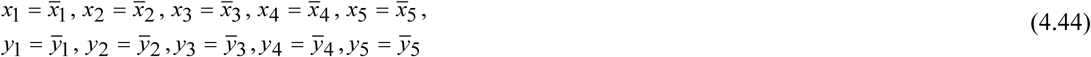

and

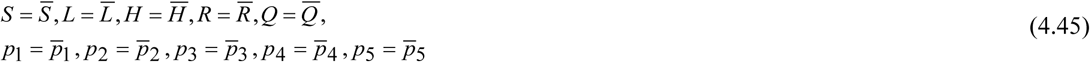

Hence the solution of (4.22) is unique for small time. So the proof is complete.

## 5. Numerical Simulations of Optimal System

In this section, a ***sensitivity analysis*** has been performed at first in order to establish which parameters have a significant influence on the reproduction number. For this analysis, the approach described in [48] has been adopted. Sensitivity analysis is the study of how the solutions change with parameters. Results of sensitivity analysis make it easy to investigate any type of dynamic model. The normalized forward sensitivity index of a variable to a parameter is the ratio of the relative change in the variable to the relative change in the parameter. When the variable is a differentiable function of the parameter, the sensitivity index may be alternatively defined using partial derivatives [48]. The basic reproduction number for the model (2.1) is

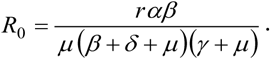

Neglecting the recruitment rate *r* and natural mortality rate *µ*, we have made a sensitivity analysis on the basic reproduction number *R*_0_ with respect to the remaining parameters *α, β, δ* and *γ*.

**Table 5.1.** Sensitivity indices of *R*_0_ to parameters

| Parameter | Description | Sensitivity index |
| --- | --- | --- |
| $\alpha$ | interaction rate among the susceptibles and light drug users | + 1.00 |
| $\beta$ | effective rate at which light users convert into heavy drug users | + 0.071085 |
| $\delta$ | removal rate from addiction without treatment | − 0.05806 |
| $\gamma$ | rate of sending heavy addicts to rehabilitation for treatment | − 0.99627 |

Parameters whose sensitivity index values are near -1 or +1 suggest that a change in the magnitudes will create a significant impact on either decreasing or increasing the size of basic reproductive number *R*_0_. From **Table 5.1**, it can easily be observed that the interaction rate parameter *α* is the most sensitive parameter to *R*_0_ as it has the highest positive sensitivity index value i.e. + 1.00. Therefore, an increase in the values of interaction rate parameter *α* will increase the size of basic reproductive number *R*_0_ which means that drug addiction will prevail for a long time. Similarly, the effective rate *β* with a positive sensitivity index value 0.071085 is also responsible for drug addiction.

From Figure 5.1, it is observed that an increase (or decrease) in the magnitude of the parameters *α* and *β* will result in an increase (or decrease) in the prevalence of drug addiction. On the other hand, the parameters *γ* and *δ* are shown to have the greatest potential to reduce the harmful situations of drug addiction when the values of these parameters are maximized. From this analysis, it is realized that the interaction rate and effective rate have a large impact on the prevalence of drug addiction. So, we should focus on these two parameters so that the harmful situation of drug addiction can be minimized. That means, we are successful to figure out that the interaction rate parameter *α* and effective rate parameter *β* are mainly liable for the harmful situation. Now we want to minimize those harmful situations by applying our three controls. For this, we have considered four scenarios. In first scenario, we have considered only the awareness and educational programs (*u*_1_). In second scenario, only the family based care (*u*_2_) is taken as control. The effectiveness of rehabilitation centers (*u*_3_) is considered as a control in third scenario. At last, all the controls are considered simultaneously in the fourth scenario. We have an intension to show that social awareness (awareness programs and family based care) is the best way to solve the situation instead of sending the addicted people to rehab centers. By this, the pressure on the rehab centers will be less and the economical cost that is required in rehab centers for treatment will be minimum. To find the answers of all questions, numerical simulations using optimal controls have been performed by using MATLAB with the set of parameter values given in **Table 5.2**. Due to the deficiency of data on drug addiction, precise adjustment is however detrimental to make. Nevertheless, some of the parameters have been assumed in some realistic point of view and the ranges are considered based on the guidance of previous literatures on drug abuse epidemics. Finally, the purpose of this simulation is to state the behaviors of our model and describe the dynamics of drug addiction.

**Figure 5.1.**
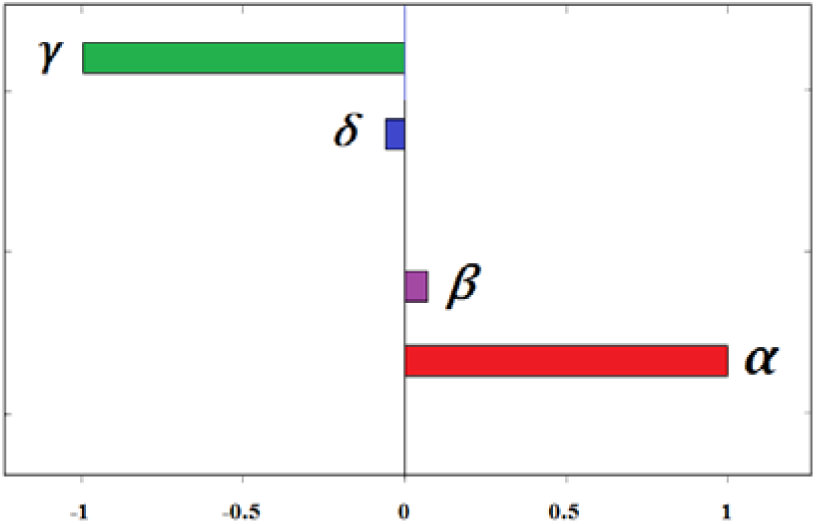
Effects of parameter variations on *R*_0_

**Table 5.2.**
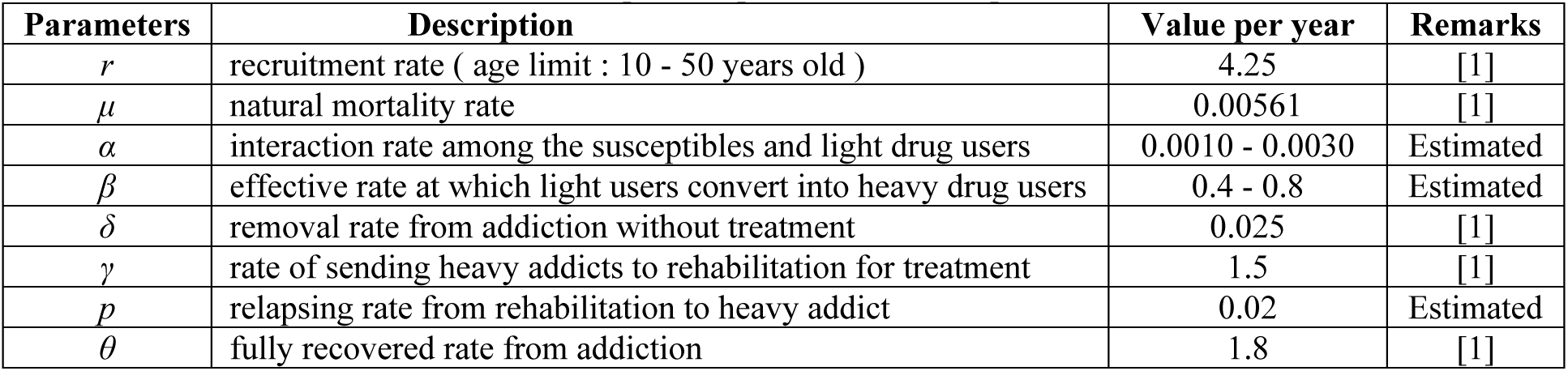
Description of parameters and respective values

| Parameters | Description | Value per year | Remarks |
| --- | --- | --- | --- |
| $r$ | recruitment rate ( age limit : 10 - 50 years old ) | 4.25 | [1] |
| $\mu$ | natural mortality rate | 0.00561 | [1] |
| $\alpha$ | interaction rate among the susceptibles and light drug users | 0.0010 - 0.0030 | Estimated |
| $\beta$ | effective rate at which light users convert into heavy drug users | 0.4 - 0.8 | Estimated |
| $\delta$ | removal rate from addiction without treatment | 0.025 | [1] |
| $\gamma$ | rate of sending heavy addicts to rehabilitation for treatment | 1.5 | [1] |
| $\rho$ | relapsing rate from rehabilitation to heavy addict | 0.02 | Estimated |
| $\theta$ | fully recovered rate from addiction | 1.8 | [1] |

### First Scenario

In this scenario, only the awareness and educational programs (*u*_1_) is considered whereas the other two controls: family based care (*u*_2_) and the effectiveness of rehabilitation centers (*u*_3_) are set to zero (i.e. *u*_1_ ≠ 0, *u*_2_ = 0, *u*_3_ = 0). Then the following figures have been obtained.

### Second Scenario

In this scenario, only the family based care (*u*_2_) is taken in absence of other two controls: awareness and educational programs (*u*_1_) and the effectiveness of rehabilitation centers (*u*_3_) are set to zero (i.e. *u*_2_ ≠ 0, *u*_1_ = 0, *u*_3_ = 0). Then the following results have been obtained.

### Third Scenario

In this scenario, we activate the control *u*_3_ (effectiveness of rehabilitation centers) whereas the other two controls: awareness and educational programs (*u*_1_) and family based care (*u*_2_) are set to zero (i.e. *u*_3_ ≠ 0, *u*_1_ = 0, *u*_2_ = 0). Then we have obtained the following figures.

### Fourth Scenario

In this scenario, all the controls *u*_1_, *u*_2_, *u*_3_ are considered simultaneously (i.e. *u*_1_, *u*_2_, *u*_3_ ≠ 0). Using all these controls combinedly, the dynamics of light addicted, heavy addicted, addicted population in rehab and quitter (recovered) population have been shown in following figures.

**Figures 5.2 - 5.17** show the dynamics of all scenarios for the optimal controls u_1_, u_2_and *u*_3_. To describe the whole phenomena in an easy manner, the following bar charts have been constructed. Since the main purpose is to minimize the number of light and heavy addicted population, all the scenarios illustrated before have been analyzed.

**Figure 5.2.**
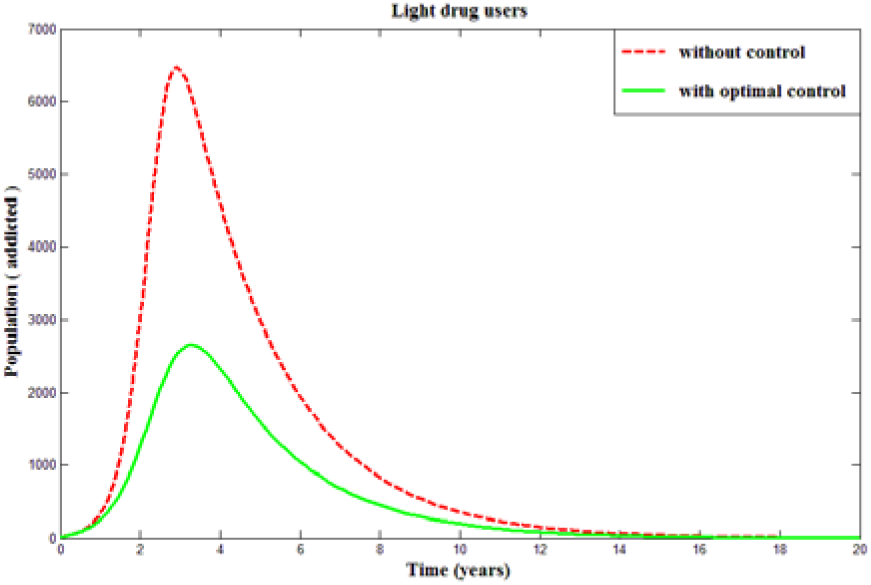
Dynamics of light addicted population using awareness and educational programs (*u*_1_) as optimal control

**Figure 5.3.**
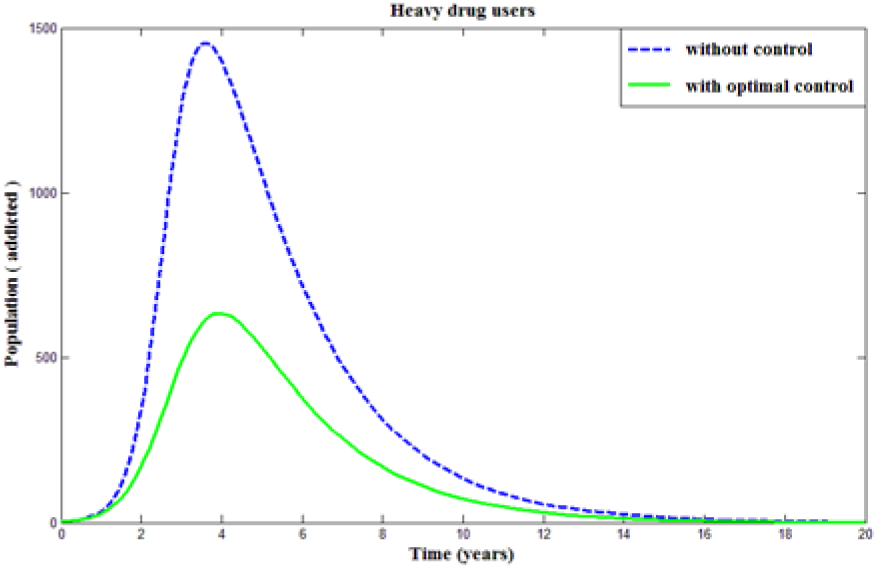
Dynamics of heavy addicted population using awareness and educational programs (*u*_1_) as optimal control

**Figure 5.4.**
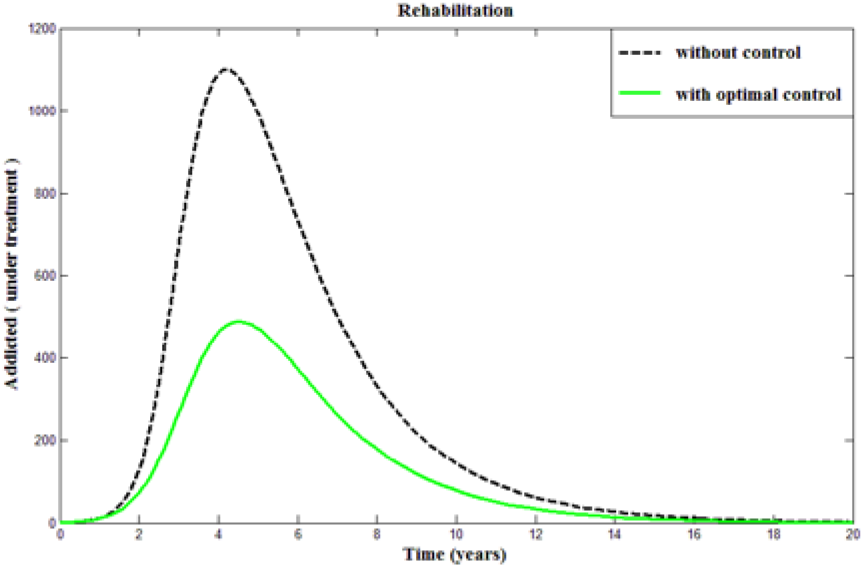
Dynamics of addicted population in rehabilitation using awareness and educational programs (*u*_1_) as optimal control

**Figure 5.5.**
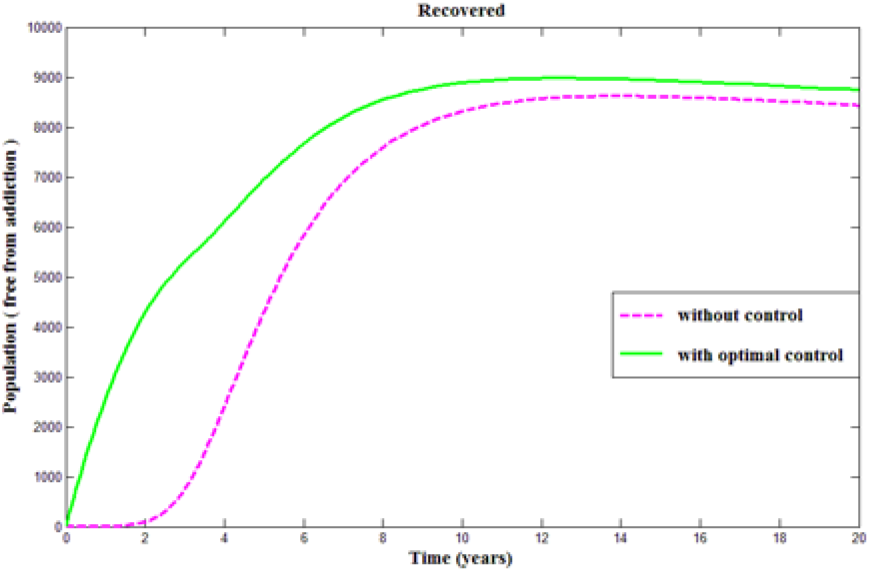
Dynamics of quitter population using awareness and educational programs (*u*_1_) as optimal control

**Figure 5.6.**
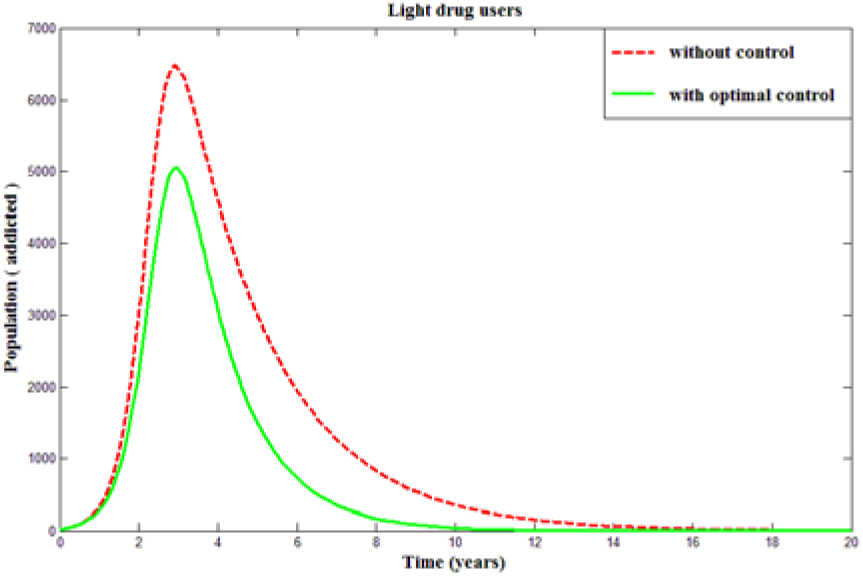
Dynamics of light addicted population using family based care (*u*_2_) as optimal control

**Figure 5.7.**
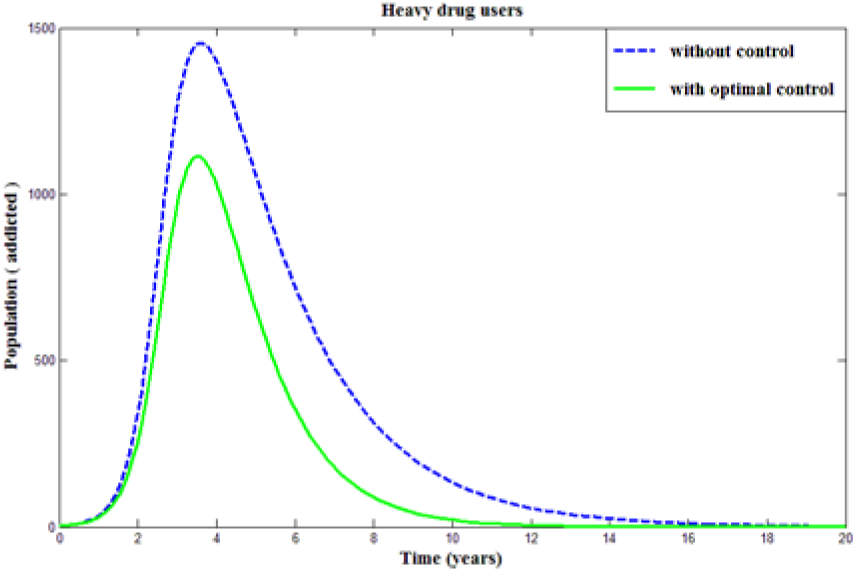
Dynamics of heavy addicted population using family based care (*u*_2_) as optimal control

**Figure 5.8.**
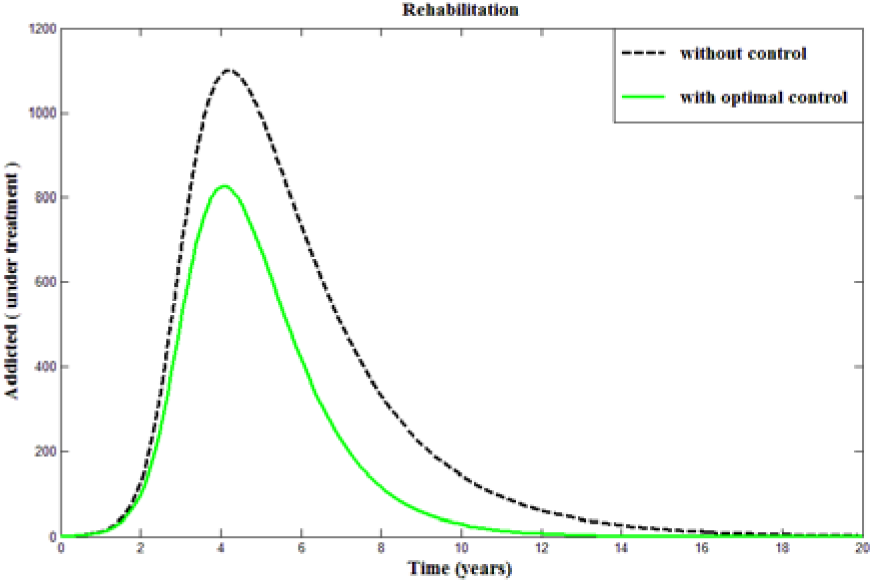
Dynamics of addicted population in rehabilitation using family based care (*u*_2_) as optimal control

**Figure 5.9.**
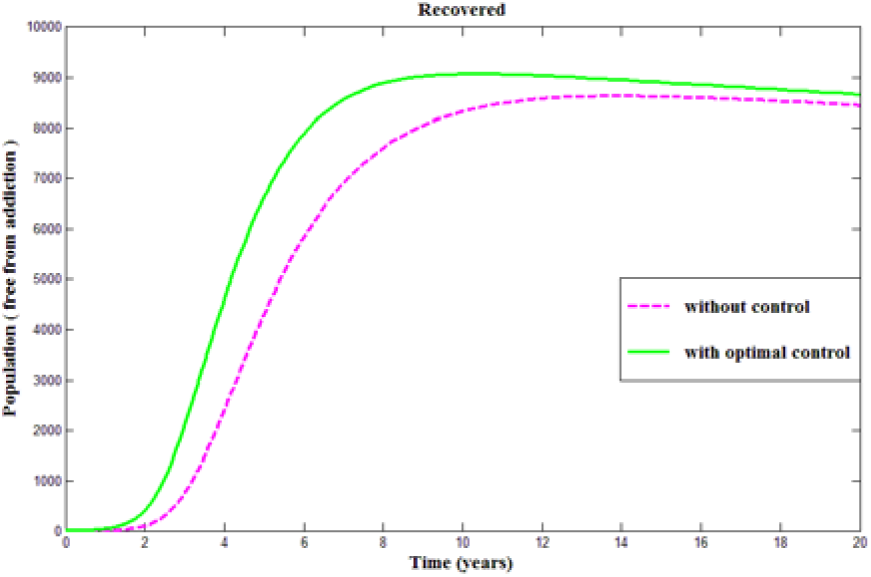
Dynamics of quitter population using family based care (*u*_2_) as optimal control

**Figure 5.10.**
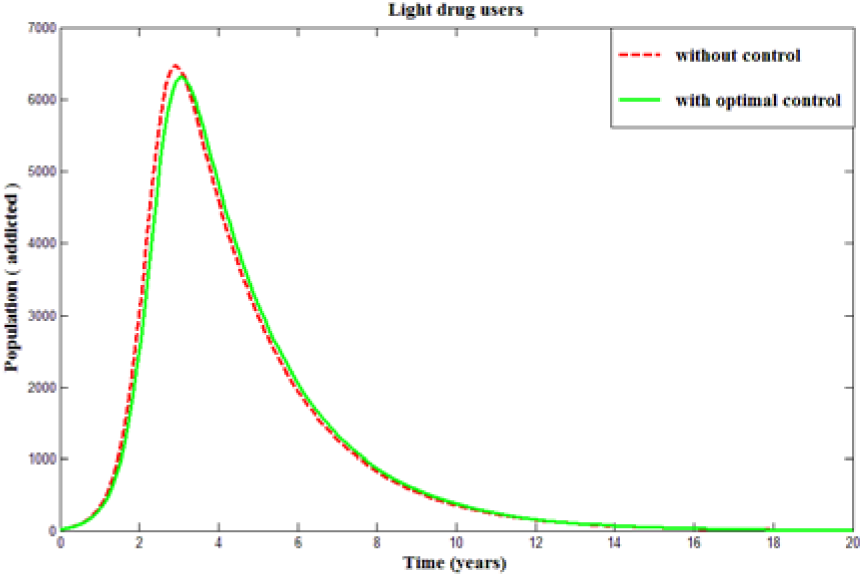
Dynamics of light addicted population using effectiveness of rehabilitation centers (*u*_3_) as optimal control

**Figure 5.11.**
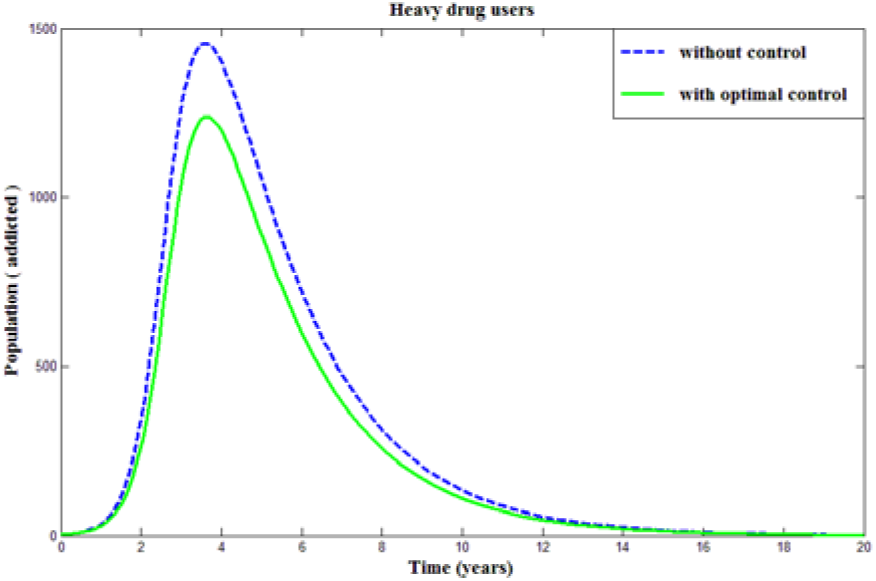
Dynamics of heavy addicted population using effectiveness of rehabilitation centers (*u*_3_) as optimal control

**Figure 5.12.**
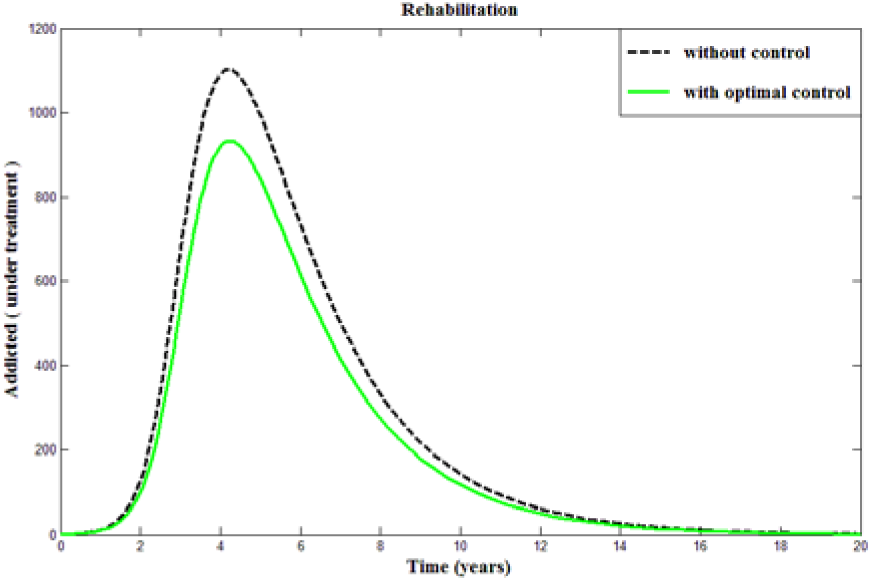
Dynamics of addicted population in rehabilitation using effectiveness of rehabilitation centers (*u*_3_) as optimal control

**Figure 5.13.**
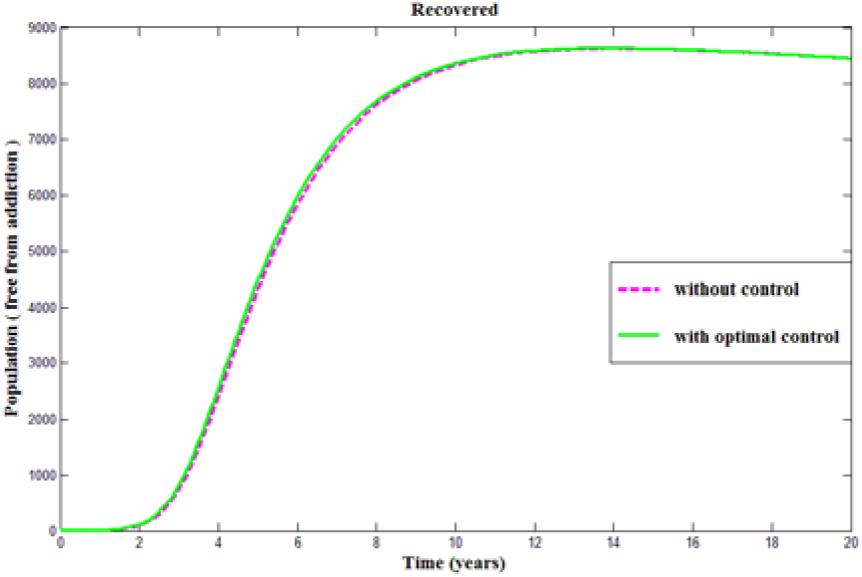
Dynamics of quitter population using effectiveness of rehabilitation centers (*u*_3_) as optimal control

**Figure 5.14.**
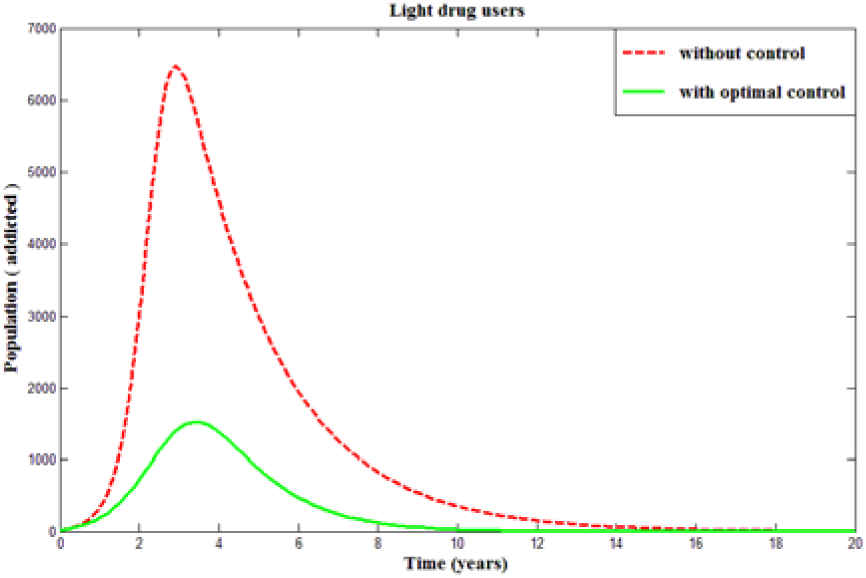
Dynamics of light addicted population using *u*_1_, *u*_2_, *u*_3_ as optimal control

**Figure 5.15.**
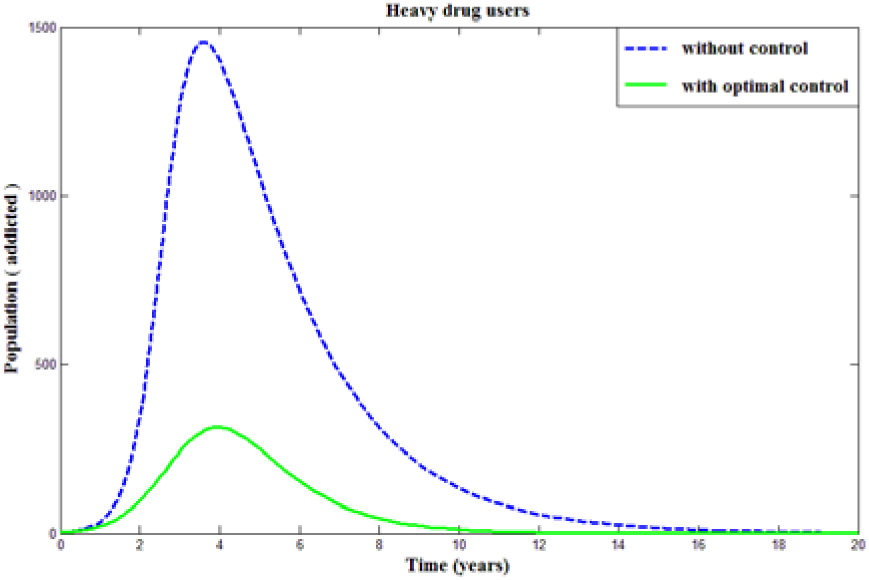
Dynamics of heavy addicted population using *u*_1_, *u*_2_, *u*_3_ as optimal control

**Figure 5.16.**
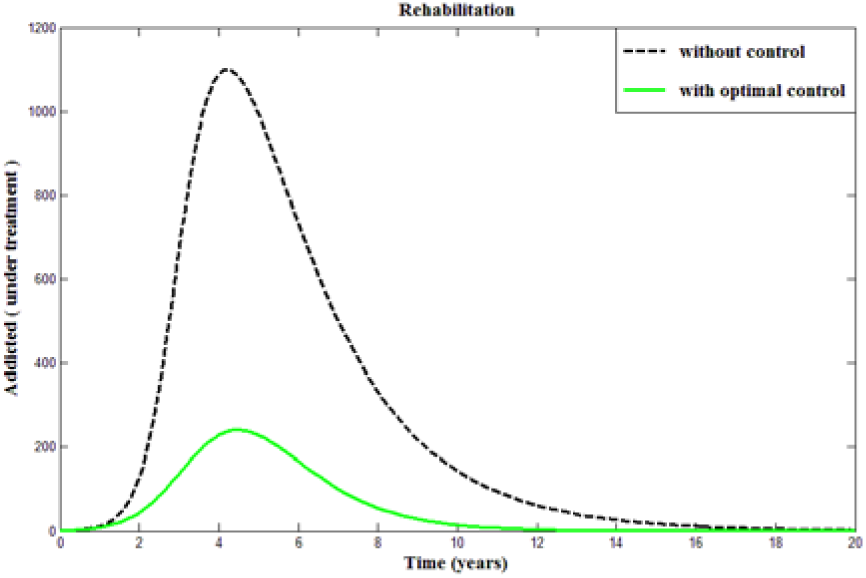
Dynamics of addicted population in rehabilitation using *u*_1_, *u*_2_, *u*_3_ as optimal control

**Figure 5.17.**
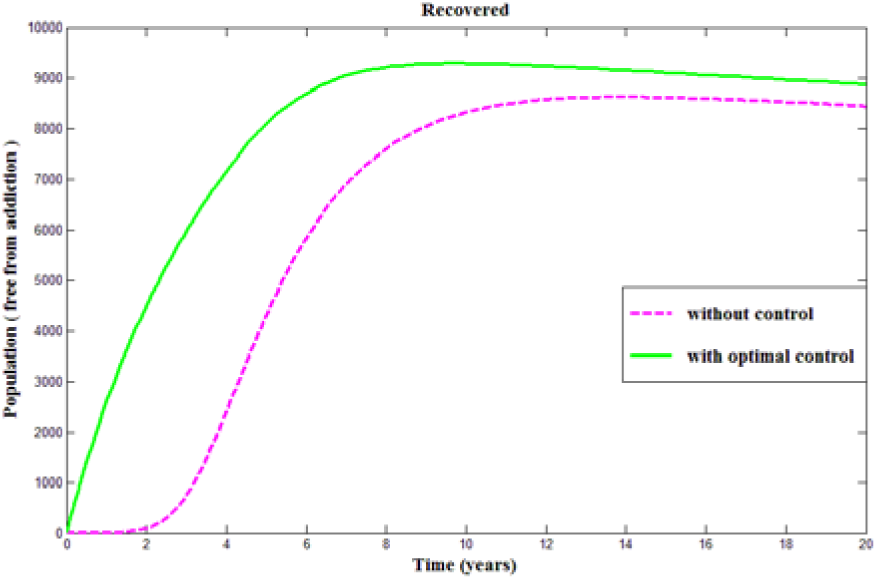
Dynamics of quitter population using *u*_1_, *u*_2_, *u*_3_ as optimal control

From **Figure 5.18**, it can be observed that the awareness and educational programs (*u*_1_) is more effective than the other two controls, when we use only one control. Moreover, family based care (*u*_2_) can also be useful because family is the place where the addicted fellows can get a better circumstance. But when we use the combination of all controls simultaneously, then it shows a highest impact in minimizing the number of light addicted population. **Figure 5.19** shows that the first scenario can be effective at the time of applying only one control i.e. the awareness and educational programs (*u*_1_) is more effective to diminish the size of heavy addicted population. But if we apply all the controls, the number of heavy addicted population can be minimized within a shortest possible time.

**Figure 5.18.**
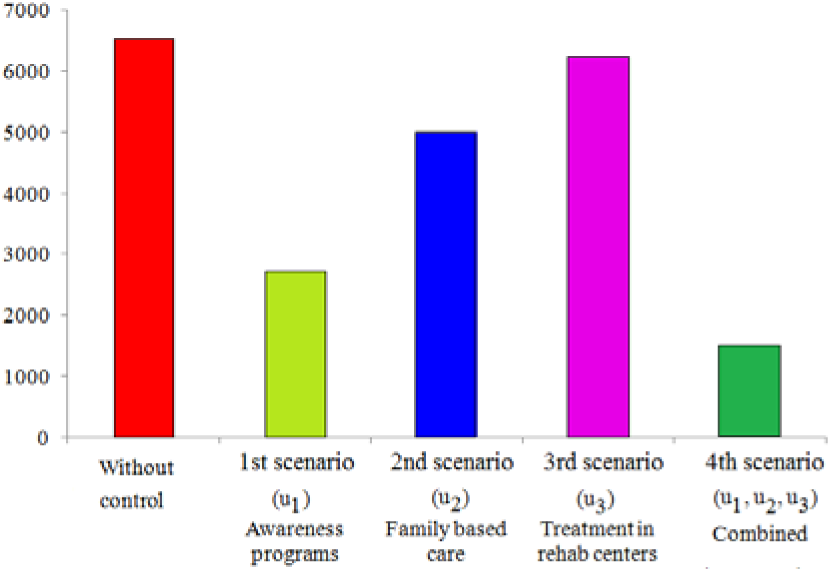
Approximate peak value of light addicted population for all scenarios

**Figure 5.19.**
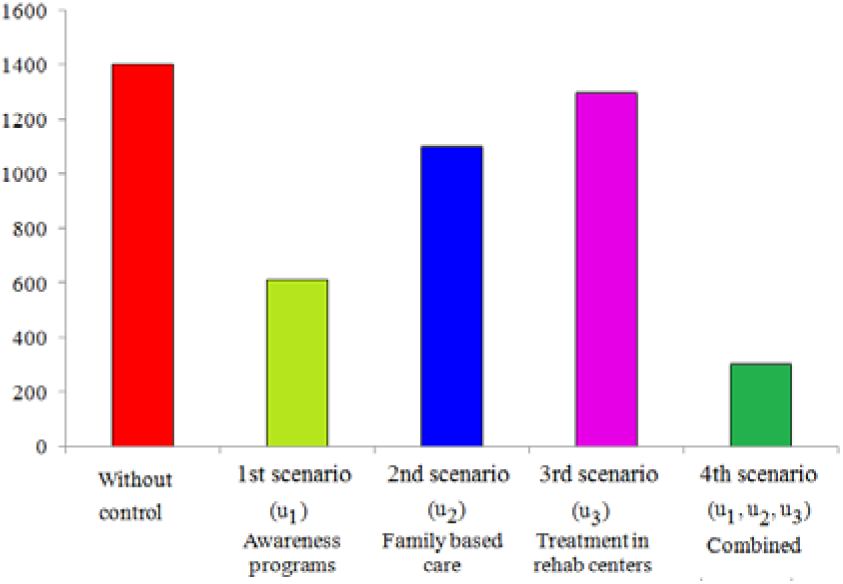
Approximate peak value of heavy addicted population for all scenarios

From **Figure 5.20**, it is observed that although treatments are available in the rehabilitation centers, these treatments (*u*_3_) are not adequate enough to minimize the number of addicted patients. So, we must have to apply the other two controls: family based care (*u*_2_), awareness and educational programs (*u*_1_) at the very early period to control the whole situation. By this, the pressure on the rehabilitation centers will be minimum and we may get a better result from the very beginning which fulfills our main desire. Moreover, it is clear that a significant change or less addicted patients can be obtained if we use all the controls *u*_1_, *u*_2_ and *u*_3_ at the same time.

**Figure 5.20.**
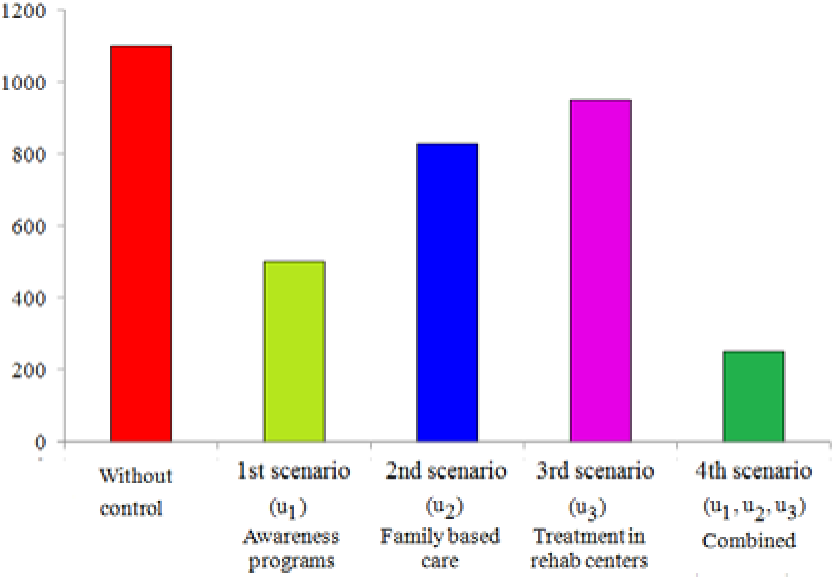
Approximate peak value of addicted population in rehab for all scenarios

**Figure 5.21** shows the number of fully recovered population after 4 years. We can see that the approximate value is about 2,000 when there is no control. Applying only one control, the size of the population increases to near about 6,000 in first scenario for *u*_1_, 4,000 in second scenario for *u*_2_ and 2,200 in third scenario for *u*_3_. So at the time of using only one control, the awareness and educational programs *u*_1_ is better than the other two controls. When all the controls *u*_1_, *u*_2_ and *u*_3_ are applied combinedly, then the recovered population increases to 7,000 in fourth scenario which is more effective.

**Figure 5.21.**
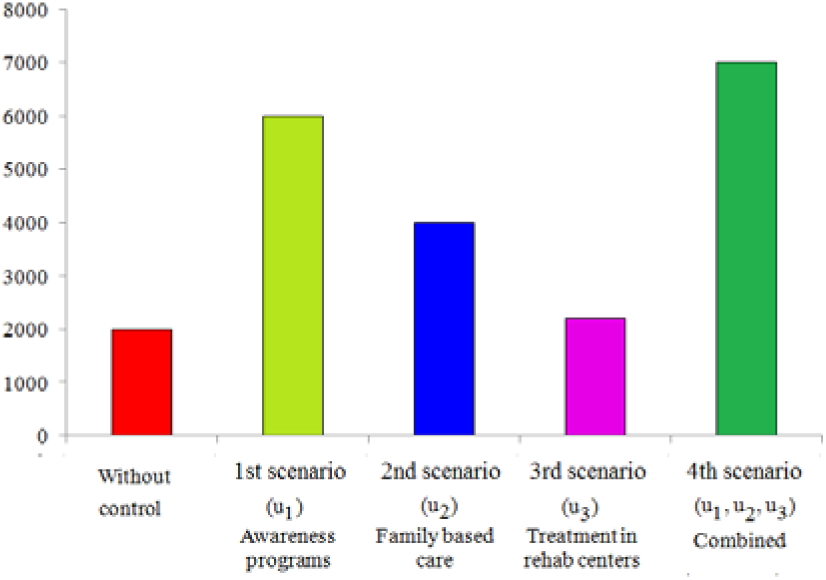
Approximate value of quitter (recovered) population after 4 years for all scenarios

The profiles of the optimal controls have been displayed in **Figure 5.22**. It is noticed that the profiles of the optimal controls *u*_1_ and u_2_ are analogous which indicates that both *u*_1_ and *u*_2_ require an initial strong start that should be continued for a greater period of time. That means, awareness and educational program with family based care should be maximized to reduce the harmful situation. On the other hand, the control profile u_3_ demonstrates similar trends. From the figure, it is revealed that supports from the rehabilitation centers should be increased gradually andffcient treatment services should be provided and maintained throughout the treatment period.

**Figure 5.22.**
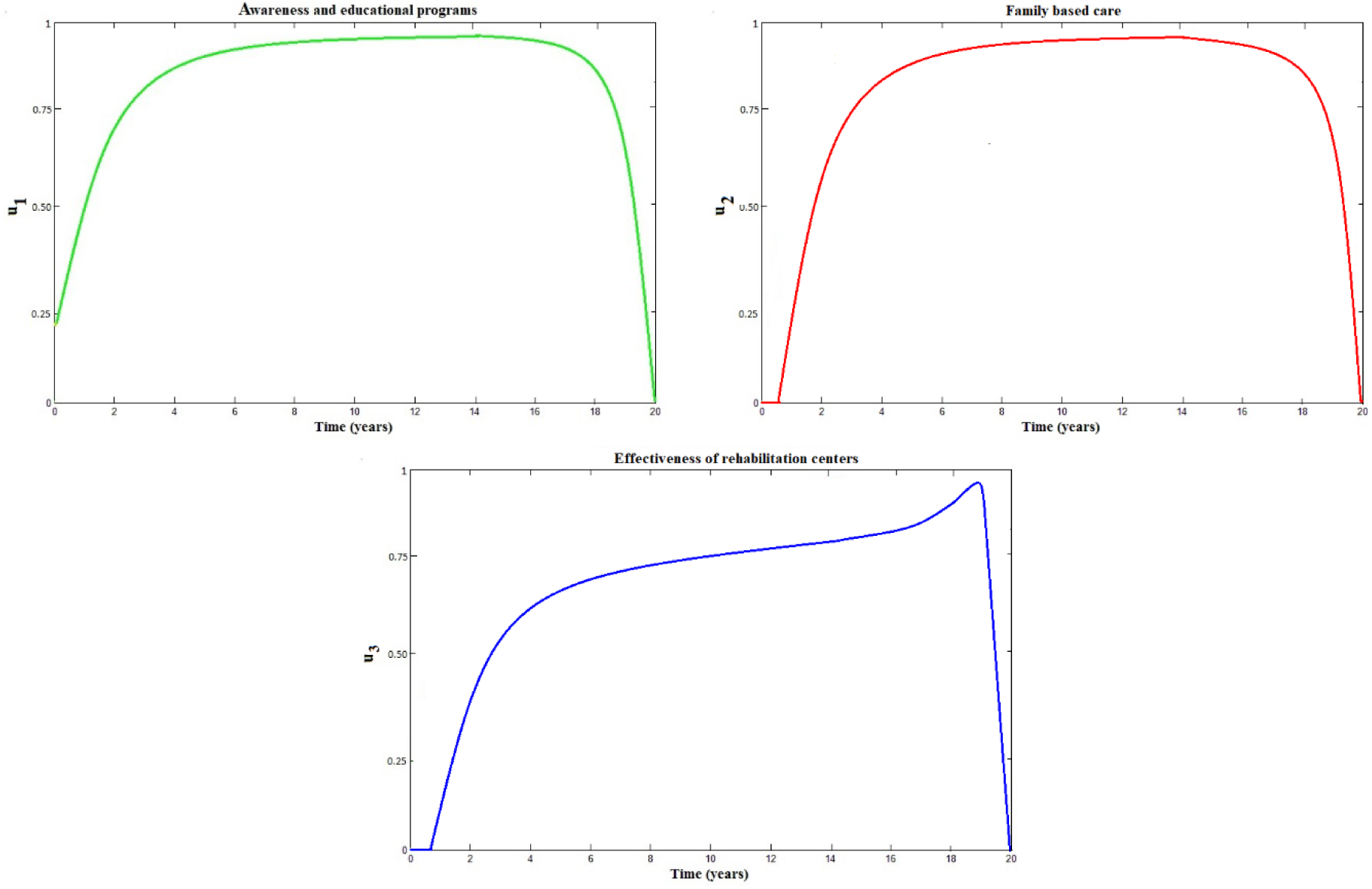
Profiles of the optimal controls *u*_1_, *u*_2_, *u*_3_ with weight parameters *A* = 0.5, *B* = 0.4 and *C* = 0.2

From the simulation results, it can be understood that the combination of educational campaigns, family care and effective treatment is very effective and plays a vital role in minimizing the harmful consequences of drug abuse in a shortest possible time compared to the case of without controls.

## 6. Conclusions

After performing analytical and numerical simulations, we have found that the control, awareness and educational programs, can be the most effective way to minimize the addicted population in a shortest possible time. The family based care is also a useful medium for controlling the adverse situation. By this, the pressure on the rehabilitation centers will be less as such institutions always work for the betterment of social problems. If the controls, family based care, awareness and educational programs, are possible to apply at the very beginning, the economical cost that is required in rehabilitation centers for treatments will be minimum. If we observe the controls in practical life, we must see that all the controls are effective from their own sides to reduce the harmful effects of drug addiction. Normally, family is the best place where the family members, especially the young generation, may find a suitable circumstance. The parents and guardians always give guidance to their lovable sons, daughters or relatives and take care of them about their studies. Such guidelines must help the young generation not to become addicted. This control works till the children are at home and in touch of their families. But every students has to go outside of their family to colleges or universities for their educational purposes. This is the most crucial time when the young generation may become addicted as they are not in the boundaries of their families. In this situation, our assumed control, awareness and educational programs, can be the most constructive way to prevent them from being addicted. Awareness and educational programs always focus on the harmful effects of taking drug substances so that the young generation may become conscious and stay away from the evil deeds. Despite taking all the initiatives, i.e. applying family based care and awareness programs, anyone may become a drug addicted person. In that situation, he or she might get a proper treatment from the rehabilitation centers. But rehabilitation centers are very rare in rural areas. Some families also feel hesitate to send their addicted relatives to the rehabilitation centers for treatment.

Moreover, the treatments in rehabilitation centers are quite expensive. This is the main reason why the effects of this control are less and it takes a long time to minimize the addicted population than the other two controls. That means, a minimum number of addicted population can be found in a shortest possible time if family based care, awareness and educational programs are applied. Moreover, the effects of drug addiction can be controlled in an effective manner if we apply all the controls at the same time.

In short, we have formulated a general model showing the phenomena of drug addiction and specified the ways about how to control it from a locality in a shortest possible time. Our model will increase consciousness among the young generation and may help the social workers to make plans regarding drug addicted population. If it is possible to apply all the preventive ways that have been shown in this model, anyone can hope for a better country which will be free from drug addiction.

## Data Availability

The data used to support the findings of this study are included within the article.

## Acknowledgment

The first author was supported by the Ministry of Science and Technology, Government of the People’s Republic of Bangladesh in his MSc program with the NST Fellowship 2017-18, Grant No. 39.00.0000.012.02.009.17-662, Serial: 128, Date-10.01.2018. This partial financial support is greatly acknowledged.

## Data Availability

The data used to support the findings of this study are included within the article.

## Authors’ Contributions

MHAB designed the study, performed the conceptualization and methodological analysis and model formulation of the first draft of the manuscript. MAII analyzed the model analytically and wrote the programming codes to perform the computational analysis and also contributed to literature searches and calculated the real data to estimate the parameters. All authors have read and agreed to the publish the final version of the manuscript.

